# Influence of mindfulness training on attention processing in individuals with chronic pain

**DOI:** 10.1101/2025.05.28.25328533

**Authors:** Michael Yufeng Wang, Neil W Bailey, Paul B Fitzgerald, Bernadette Mary Fitzgibbon

## Abstract

**Objective:** The dynamic impact of chronic pain on attention can lead to impairments in overall cognitive functioning. Mindfulness is a practice often adopted for pain management that includes elements of attention training. It is thought to improve the experiential acceptance of pain, thus reducing pain-sensitivity and pain-related attentional biases. The aim of this study was to examine whether experience with mindfulness is associated with altered attentional functioning in participants with chronic pain.

**Methods:** We collected an online sample of 128 participants across four groups: 31 individuals with chronic pain who practice mindfulness (Pain-Meditators), 29 individuals with chronic pain who have not practiced mindfulness (PainNon-Meditators), 32 healthy individuals who practice mindfulness, and 36 healthy individuals who do not practice mindfulness. General attentional functioning was measured using the using the Short-Attention Network Task (short-ANT) and attention bias to pain was measured using the Pain Dot-Probe task.

**Results:** An overall effect in reaction time (RT) was found across both (all *p* <.05), with post-hoc analyses finding that Pain-Meditators reacted faster on both the short-ANT (*p* = 0.014) and dot-probe attention tasks (p = < 0.001) compared to PainNon-meditators. PainNon-meditators showed higher alerting network scores on the short-ANT than all other groups (*p* = 0.005), indicating more reliance on cues to alert attention. No differences were found between the pain groups in attentional bias to pain-related words (*p* = 0.84).

**Conclusion:** The results provide evidence that mindfulness practice is associated with altered attention performance in individuals with chronic pain and may mitigate the effects of chronic pain on attention functioning. Our results provide some of the only experimental research to investigate the effects of mindfulness training on attention processing in individuals with chronic pain.

## Introduction

Pain is characterised by the International Association for the Study of Pain (IASP) as an unpleasant experience associated with existing or possible injury (Merskey & Bogduk, 2011). Pain is believed to play an adaptive role in injury prevention and harm minimisation and is thus thought to be prioritised for attention processing (Eccleston, 1994; Eccleston & Crombez, 1999; Garland, 2012; Woolf, 2010). However, in conditions where pain is chronic (i.e., ongoing pain without relief), attention to pain stimuli can further accentuate and heighten the perceived threat of pain. Thus, the relationship between pain and attention is considered bidirectional, whereby attention toward pain can heighten perceived threat and further drive attention toward the site of pain (Arntz & De Jong, 1993; Eccleston et al., 1997; Gong et al., 2019; Moore et al., 2012).

For individuals living with chronic pain, this relationship between pain and attention can therefore be detrimental. Specifically, not only can increased pain attention enhance the pain experience, but difficulty orienting attention away from pain to focus on task-relevant information, can impact broader cognitive functions important for daily tasks such as executive functioning (Keogh et al., 2013; Moore et al., 2012). Furthermore, reduced top-down attention control to direct attention away from pain has been shown to heighten pain vigilance, and evidence have linked greater pain vigilance to higher ratings of pain intensity and pain-related anxiety, both contributing to the maintenance of negative pain experiences and may therefore contribute to the perpetuation of chronicity (Arntz & De Jong, 1993; Mogg & Bradley, 2016).

Modulating attention may thus be an effective form of pain management for chronic pain (Elomaa et al., 2009; Morley et al., 2006). One intervention that is already used in pain management and is thought to influence attention is mindfulness (Curtin & Norris, 2017; McClintock et al., 2019; Veehof et al., 2016). Mindfulness trains top-down self-regulation of attention and promotes the experiential acceptance of internal and external experiences (Bishop et al., 2004). In pain management, mindfulness-related interventions have been shown to enhance attentional control with evidence of greater ability to disengage from pain-related cues in individuals with chronic pain who practice mindfulness (Elomaa et al., 2009; Garland & Howard, 2013; Majeed et al., 2018). Furthermore, mindfulness training has been demonstrated to lower pain vigilance and pain-related anxiety which can alleviate the impact of pain on attention processes (Cramer et al., 2012; Curtin & Norris, 2017; Majeed et al., 2018).

While mindfulness practice is believed to reduce the impact of pain on attention-related processes (Tsur et al., 2021), to our knowledge, no behavioural studies have investigated associations between mindfulness practice and general attentional functioning in individuals with chronic pain. The primary aim of the present study was therefore to examine whether mindfulness training influences attention processing in individuals with chronic pain. Investigating the effects of mindfulness training on attention functioning in individuals with chronic pain will provide a better understanding of the mechanism by which mindfulness can modulate pain, and thus support its use in pain management.

## Methods

### Study Design

The present study design tested three central aims. First, we examined the effects of mindfulness practice on general attention functioning and attentional bias to pain in two chronic pain groups; individuals with chronic pain who practice mindfulness (referred to as Pain-Meditators for brevity in the rest of this manuscript) and individuals with chronic pain who do not practice mindfulness (referred to as PainNon-Meditators in this manuscript). Secondly, we assessed whether the potential protective effects of mindfulness on attention processing in the Pain-Meditator group enabled attention task performance comparable to the general population who do not practice mindfulness (Non-meditators) as well as the general population who practice mindfulness (Meditators). Thus, we used a 4x2 mixed between-within subjects design (Pain-Meditators, PainNon-Meditators, Meditators, and Non-meditators) with independent variables being task performance on two selected attention tasks that measured; 1) general attention functioning and 2) attentional bias to pain-relate cues. Lastly, a regression analysis was used to investigate whether attention performance in the Pain-Meditator group was related to self-reported levels of pain resilience, mindfulness, and pain-vigilance.

### Participants

A total of 176 participants took part in the study. Forty-eight participants were excluded from the analyses due to incomplete surveys and/or cognitive tasks. The final sample consisted of 128 participants across four groups: 31 Pain-Meditators, 29 PainNon-Meditators, 32 Meditators, and 36 Non-meditators. The ages of participants ranged from 21 to 61 (*M* = 38.77, *SD* = 10.65) and did not differ significantly between groups (*F*[3, 124] = 2.11, *p* = 0.1). Diverse gender representation options were provided to participants in line with best practice recommendations for collecting gender data (CFA San Diego, 2022; National LGBT Health Education Center, 2015). In terms of responses for gender, 59 participants identified as male, 69 participants identified as female, and one participant identified as non-binary. As only one participant identified as non-binary, the non-binary participant was removed from statistical analyses that assessed gender (due to the lack of statistical power for the creation of a non-binary gender group) but included in all other analyses. No significant differences were found between the Pain-Meditators, PainNon-Meditators, Meditators, and Non-meditators groups in gender (*X*^2^[6] = 5.61, *p* = 0.49) or education level (*X*^2^[9] = 10.92, *p* = 0.28). Participants were recruited online via ads on social media and topic-relevant forums (e.g., mindfulness, chronic pain), as well as in-person through pain management centres.

Selection criteria for participants in both chronic pain groups were identified by self-report symptoms as defined by the International Association for the Study of Pain; persistent pain beyond three months or longer without biological value (Merskey & Bogduk, 2011). Information was also collected for each participant about the type of pain, length of pain, and location of pain etc., (see questionnaire section below for a detailed summary). Both the Pain-Meditator and Meditator groups were identified by self-reports of mindfulness experience. Inclusion criteria for both the Pain-Meditator and Meditator groups were: 1) that the mindfulness practice included a component of either focused attention or body scan, and 2) a component of their practice followed Kabat-Zinn’s (1994) definition of mindfulness-meditation - “*paying attention in a particular way: on purpose, in the present moment, and nonjudgmentally* ”. Both meditator group participants were included in the study if their self-reported description of their practice met the inclusion criteria, as determined by a trained mindfulness researcher (MW). Both PainNon-Meditators and Non-meditators were required to have less than two hours of lifetime meditation experience.

## Materials

### Questionnaires

All participants completed a demographic questionnaire along with the Generalized Anxiety Disorder-7 (GAD-7; Spitzer et al., 2006), Patient Health Questionnaire (PHQ-9; Kroenke et al., 2001), the Mindful Attention Awareness Scale (MAAS; Brown & Ryan, 2003), and the Pain Vigilance and Awareness Questionnaire (PVAQ; McCracken, 1997). For both the Pain-Meditator and Meditator groups, participants completed an additional Mindfulness Experience Questionnaire (questions are provided in the supplementary materials). Participants who self-identified as having a diagnosis of a chronic pain condition (Pain-Meditators and PainNon-Meditators) completed an additional questionnaire to characterise their pain experience (e.g., length of pain, location of pain – the full list of questions are provided in the supplementary materials) as well as the Pain Resilience Scale (Ankawi et al., 2017).

#### Generalized Anxiety Disorder-7 (GAD-7; Spitzer et al., 2006)

The GAD-7 is a seven-item scale designed to screen for generalised anxiety disorder. Spitzer et al. (2006) reported Cronbach α = 0.92, indicating high internal consistency. Scores range from 0 to 21 with higher scores suggesting a greater indication of generalised anxiety disorder and anxiety-related impairment.

#### Patient Health Questionnaire (PHQ-9; Kroenke et al., 2001)

The PHQ-9 is the depression module of the Patient Health Questionnaire used to screen depressive symptoms according to the DSM-IV (Kroenke et al., 2001). The scale consists of nine items with scores ranging from 0 to 27. Internal reliability was reported by Kroenke et al. (2001) as Cronbach α = 0.89, suggesting good internal consistency.

#### The Mindful Attention Awareness Scale (MAAS; Brown & Ryan, 2003)

The MAAS is a 15-item self-report scale designed to measure dispositional mindfulness. Particularly, the MASS measures experiential acceptance – i.e., attending to experiences without judgement, observing but not reacting (Brown & Ryan, 2003). Participants responded to statements such as “*I find it difficult to stay focused on what’s happening in the present* ” on a 6-point Likert scale ranging from 1 (almost always) to 6 (almost never), where high scores reflect higher dispositional mindfulness and experiential acceptance. The Cronbach α as reported by Brown and Ryan (2003) was 0.89 suggesting good internal consistency.

#### Pain Vigilance and Awareness Questionnaire (PVAQM; cCracken, 1997)

The PVAQ is a 16-item questionnaire measuring two subcomponents of attention to pain; 1) attention to pain (“*I focus on feelings of pain*”) and 2) vigilance to changes in pain (“*I am aware of quick changes in pain*”; McCracken, 1997). The items are rated on a 6-point Likert scale ranging from 0 (never) to 6 (always). Good internal consistency was reported among chronic pain populations; attention to pain subscale was reported as Cronbach α 0.85, attention to changes in pain subscale was reported as Cronbach α 0.80, and the overall Cronbach α was 0.83 (Roelofs et al., 2003).

#### Pain Resilience Scale (PRS; Ankawi et al., 2017)

The PRS is a measure of behavioural perseverance and cognitive/affective positivity concerning pain. It consists of 14-items rated on a 5-point Likert-type scale (0 = not at all, 4 = all the time). Scores range from 0 to 56, with higher scores indicating greater pain resilience. Cronbach’s alpha reported for the overall scale is α = 0.92, and Cronbach’s alpha reported for the two subscales were: 1) behavioural perseverance α = 0.90 and 2) cognitive/affective positivity α = 0.91 (Ankawi et al., 2017).

#### Qualitative Measures

The Mindfulness Experience Questionnaire was developed for this study to assess the practice history, frequency, and mindfulness style of participants who self-identified as meditators with and without a diagnosis of chronic pain. The questionnaires contained both multiple choice and short answer questions such as “*On average how many minutes per day do you typically meditate for*?” and “*Please list all meditation styles and shifts in your frequency of practice over the years* ”.

This questionnaire was developed based on the methods by Hasenkamp and Barsalou (2012), which estimates lifetime meditation experience. The Pain Characterisation Questionnaire was developed for this study to assess the length of pain, location of pain, and intensity of pain in participants who self-identified as having a diagnosis of chronic pain. The questionnaire contained questions (both text response and multiple choice) such as “*How did your pain begin*” and “*How long has your pain been present*”. Both questionnaires are presented in Supplementary Materials 1.

### Behavioural Tasks

In addition to the above questionnaire, all participants completed two attention-related cognitive tasks to measure general attention functioning (the short-Attention Network Task) and pain-related attentional bias (pain-related dot-probe).

#### Short-Attention Network Task

The short Attention Network Task (short-ANT) is a 10-minute version of the original Attention Network Task (which takes 20 minutes) designed to measure three attention networks (alerting, orienting, and executive control; Fan et al., 2002). Response time (RT) for all three attention networks measured using the short-ANT have shown a strong correlation with the original version (Weaver et al., 2013). On each trial, participants were presented with a fixation cross (400ms) followed by a cue (*) for 100ms. After the cue phase, five arrows are presented for 1700ms either above or below the fixation cross. The cue can either be presented in the centre of the screen, in the same location as the centre arrow in the upcoming five arrows (spatial cue), or not shown at all.

Participants were asked to select the direction of the middle arrow as quickly as possible. The other arrows act as distractors and were presented as pointing in the same or opposite direction as the centre arrow. Thus, the short-ANT has two flanker conditions (congruent, incongruent) and three cue conditions (centre cue, no cue, and spatial cue). The average RT to correct responses was calculated for each condition as well as the overall RT across all trials, providing a measure of overall attention efficiency. Attention network index scores (alerting, orienting, and executive control) as measured by the short-ANT are calculated using differences in RT for correct responses between conditions (Fan et al., 2002, 2005). The alerting network was assessed by subtracting the mean RT to the centre cue condition from the mean RT to the no cue condition. Orienting network performance was measured by subtracting the mean RT to the spatial cue condition from the mean RT to the centre cue condition. Lastly, executive control was measured by subtracting the mean RT to congruent flankers from the mean RT of incongruent flankers. The short-ANT consists of 12 practice trials and 120 experimental trials. D-prime (*d’*) scores were calculated to assess the overall performance accuracy (correct responses) of participants. When hits and false alarms were 1 and 0 (indicating all correct) they were replaced with 0.99 and 0.01 to determine an approximation of *d’*. A higher *d’* indicates that participants performed with fewer misses and false alarms, and therefore were more accurate.

#### Pain related Dot-Probe

The dot-probe task was used to assess attentional bias to pain-related stimuli. Participants first attended to a fixation cross in the centre of the screen lasting 500ms. Two words were then presented for 500ms, with one word presented above and one below the fixation cross. The words consisted of one neutral word (i.e., plastic) and one pain-related word (i.e., aching). The words were followed by a dot-probe (**·**) in the same location as one of the words. The dot-probe lasted 1000ms during which participants responded to the location of this dot probe, pressing one button if the probe was in the upper half of the screen and another if it was in the lower half of the screen. Thus, there are two factors and four conditions for the task: target word (pain vs neutral) x congruence (congruent vs incongruent). Participants completed a total of 10 practice trials and 100 experimental trials. The dot-probe task also provides an index for attentional bias. The abbreviations represented in the formula to calculate attention bias index are as follows; t is the target word (the pain related word), d is the probe presented on screen, u and l represent positions on the screen (upper and lower respectively). For example, tudl is when the target word is in the upper half of the screen and the dot probe is in the bottom half of the screen. Given these abbreviations, the formula for calculating the pain bias index was to insert the mean reaction time from each of the four conditions into the formula: ((tudl-tldl) + (tldu-tudu))/2 (Asmundson et al., 2005; Roelofs et al., 2002). Positive scores from this calculation reflect attentional bias to the target word, with higher numbers indicating greater attentional focus and more difficulty disengaging from the target word. *d’* scores were also calculated to assess performance accuracy.

To reduce the influence of outliers for RT calculations, all trials (across both attention tasks) with RT values more than two standard deviations above or below the individual mean within each condition for each individual participant were removed (Ratcliff, 1993).

### Procedure

The questionnaire data was collected via the Qualtrics software (Version, 2022, Qualtrics, Provo, UT), an online survey instrument. The questionnaire contained a plain language statement, which detailed the purpose of the study. Once participants consented to continue with the study, they were presented with the questionnaire in the following order: demographic questions, GAD-7, PHQ-9, MAAS, PVAQ, and the PRS, followed by mindfulness and chronic pain specific questionnaires. Once participants completed the questionnaire and consented to continue further, another link was provided to download the computer software Inquist (version 6.3.0; 2020) which participants used to complete the short-ANT and dot-probe tasks.

### Statistical Comparisons

All analyses were conducted using the software JASP (Version 0.14.1; JASP Team, 2021). Preliminary analyses using analysis of variance (ANOVA) were conducted to examine group differences in key demographic characteristics such as age, affective variables (GAD-7 and PHQ-9), the MAAS, and pain-related measures (PVAQ and PRS). The Chi-square test was used to examine group differences in gender and education level. Due to limited statistical power for a non-binary gender group, comparisons that involved gender were conducted including two gender groups (participants who identified as males or females). As mentioned earlier, the participant who identified as non-binary was included in all other analyses.

For the primary analyses, ANOVAs were performed to assess group differences in both tasks and conditions. First, comparisons of overall RT and *d*’ for both cognitive tasks were made between all groups (Pain-Meditators, PainNon-Meditators, Meditators, and Non-meditators) to assess general attention efficiency and attentional bias to pain-relate cues. This was then followed by short-ANT specific analysis. ANOVAs were conducted to compare differences between all groups in each attention network measured by the short-ANT, which addressed our first and second aim to assess the effects of mindfulness practice on attention functioning in the context of chronic pain. A repeated measures ANOVA was also used to test whether the pain groups differed in their RT to dot-probe conditions, and a *t*-test was conducted to examine differences in pain-bias index scores, again addressing our first and second aim. Lastly, a regression analysis was conducted within the Pain-Meditators group to examine the relationship between attention performance and levels of pain resilience, mindfulness, and pain-vigilance. This addressed our third aim to determine whether attention performance in the Pain-Meditator group was related to higher levels of pain resilience, mindfulness, and lower levels of pain-vigilance. To control for multiple comparisons and the discovery of false positives, the Benjamini and Hochberg false discovery rate (FDR; Benjamini & Hochberg, 1995) was used across all analyses.

## Results

### Demographic characteristics

There were no significant differences in age, gender, or education between all groups (Pain-Meditators, PainNon-Meditators, Meditators, Non-meditators) (all *p* > 0.05). In alignment with the expected effects of pain on mood, ANOVA results revealed significant differences in mood measures across the four groups; GAD-7 = *F*(3, 124) = 6.03, *p* < 0.001, η^2^ = 0.13, and PHQ-9 = *F*(3, 124) = 10.15, *p* < 0.001, η^2^ = 0.2. Post-hoc comparisons using Tukey HSD test found no differences between pain groups (Pain-Meditators and PainNon-Meditators) in mood measures (GAD-7 and PHQ-9, all *p* > 0.05) and thus mood scores did not impact the comparisons between pain groups. Compared to the general population, PainNon-Meditators scored significantly higher on both GAD-7 and PHQ-9 than Meditators (*p* = 0.031 and *p* < 0.001 respectively) and Non-meditators (*p* = 0.007 and *p* = 0.002 respectively). Pain-Meditators also scored significantly higher than Non-meditators on the GAD-7 (*p* = 0.015) and significantly higher than Meditators on the PHQ-9 (*p* = 0.011). The remaining comparisons between groups and mood measures did not show any significant differences (all *p* > 0.05). Differences between groups in mood measures could impact the results as poor mood can impair attentional performance (Cohen et al., 2001; Eysenck et al., 2007), thus mood measures were included as covariates for analyses between all four groups. For Mindfulness (MASS) and pain measures (PVAQ and PRS), scores differed significantly between pain groups, with the PainNon-Meditator group scoring significantly higher than the Pain-Meditator group on both the PVAQ (*p* = 0.006) and PRS (*p* < 0.001), and lower on the MASS (*p* < 0.001). Scores for these measures were also significantly different in comparisons that included all groups (MAAS and pain scores were expected to differ between groups and thus were not considered confounding variables). See demographic statistics of participants presented in Table 1 below.

**Table 1.** Demographic characteristics of participants.

|  | Pain-Non-Meditators | Pain-Meditators | Meditators | Non-meditators | Statistics |
| --- | --- | --- | --- | --- | --- |
| *N* | 29 | 31 | 32 | 36 |  |
| Gender (F/M) | 12/17 | 13/17/1nb | 19/13 | 18/18 | $\chi^2(6) = 5.61, p = 0.49$ |
| Education | 12HS, 6V/T, 11BD, 0PD | 12HS, 5V/T, 12BD, 1PD | 6HS, 4V/T, 19BD, 3PD | 11HS, 9V/T, 15BD, 1PD | $\chi^2(9) = 10.92, p = 0.28$ |
|  | *M (SE)* | *M (SE)* | *M (SE)* | *M (SE)* | Statistics |
| Age | 41.07 (1.71) | 38.39 (1.8) | 35.09 (1.92) | 40.50 (1.93) | $F(3, 124) = 2.11, p = 0.1$ |
| GAD-7 | 8.55 (0.76) | 7.58 (0.53) | 5.59 (0.71) | 5.58 (0.39) | $F(3, 124) = 6.03, p < 0.001, \eta^2 = 0.13$ |
| *Post-hoc Group comparisons* | *\*\* Meditators*<br>*\*\* Non-meditators* |  | *\*\*\* Pain-Non-Meditator* | *\*\*\* Pain-Non-Meditator* |  |
| PHQ-9 | 10.66 (0.69) | 8.71 (0.69) | 5.91 (0.56) | 7.31 (0.56) | $F(3, 124) = 10.15, p < 0.001, \eta^2 = 0.2$ |
| *Post-hoc Group comparisons* | *\*\*\* Meditators*<br>*\*\* Non-meditators* | *\* Meditators* | *\*\*\* Pain-Non-Meditator*<br>*\* Pain-Meditator* | *\*\*\* Pain-Non-Meditator* |  |
| MAAS | 3.18 (0.11) | 4.59 (0.08) | 4.6 (0.14) | 3.53 (0.13) | $F(3, 124) = 35.74, p < 0.001, \eta^2 = 0.46$ |
| *Post-hoc Group comparisons* | *\*\*\* Pain-Meditators*<br>*\*\*\* Meditators* | *\*\*\* Non-meditators*<br>*\*\*\* Pain-Non-Meditator* | *\*\*\* Non-meditators*<br>*\*\*\* Pain-Non-Meditator* | *\*\*\* Pain-Non-Meditator*<br>*\*\*\* Pain-Meditators* |  |
| PVAQ | 51.35 (2) | 39.77 (2.69) | 36.34 (1.62) | 41.08 (1.69) | $F(3, 124) = 9.55, p < 0.001, \eta^2 = 0.19$ |
| *Post-hoc Group comparisons* | *\*\*\* Pain-Meditators*<br>*\*\*\* Meditators*<br>*\*\* Non-meditators* | *\*\*\* Pain-Non-Meditator* | *\*\*\* Pain-Non-Meditator* | *\*\*\* Pain-Non-Meditator* |  |
| PRS | 36.55 (2.49) | 18.48 (1.62) | 23.13 (1.51) | 25.31 (1.64) | $F(3, 124) = 16.58, p < 0.001, \eta^2 = 0.29$ |
| *Post-hoc Group comparisons* | *\*\*\* Pain-Meditators*<br>*\*\*\* Meditators*<br>*\*\*\* Non-meditators* | *\* Non-meditators*<br>*\*\*\* Pain-Non-Meditator* | *\*\*\* Pain-Non-Meditator* | *\*\*\* Pain-Non-Meditator*<br>*\* Pain-Meditators* |  |
| Meditation Experience | - | 322.13 (93.66) | 1151.90 (442.61) | - | $t(61) = 1.81, p = 0.08$ |

*Note:* $N$ = number of participants; HS = Highschool or equivalent, V/T = Vocational/TAFE, BD = Bachelor's Degree, PD = Postgraduate Degree; GAD-7 = Generalized Anxiety Disorder-7, PHQ-9 = Patient Health Questionnaire; MASS = The Mindful Attention Awareness Scale, PVAQ = Pain Vigilance and Awareness Questionnaire, PRS = Pain Resilience Scale; F = Female, M = Male, nb = non-binary; \*\*\* $p < 0.001$ , \*\* $p < 0.01$ , \* $p < 0.05$ .

### General attention processing and task accuracy

ANOVA comparison between all groups (Meditator, Non-meditator, Pain-Meditators, and PainNon-Meditators) revealed a significant difference in overall average RT on the short-ANT *F*(3, 124) = 9.77, *p* < 0.001, FDR-*p* = 0.004, η^2^ = 0.19. In post-hoc comparisons using Tukey HSD test, the PainNon-Meditator (RT *M* = 622.94) group reacted slower overall on the short-ANT than all other groups; Pain-Meditator group (RT *M* = 534.15, 95% C.I. = [-164.26, -13.32], *p* = 0.014), Meditators (RT *M* = 467.50, 95% C.I. = [-230.33, -80.54], *p* < 0.001), and Non-meditators (RT *M* = 540.72, 95% C.I. = [-155.12, -9.33], *p* = 0.02). Meditators reacted faster than the Non-meditator group (95% C.I. = [-144.19, -2.24], *p* = 0.04). The remaining comparisons between groups in the overall RT on the short-ANT task did not reveal significant differences (all *p* > 0.05). To determine the influence of mood measures (GAD-7 and PHQ-9) on group differences in overall short-ANT RT, follow-up analysis of covariance (ANCOVA) was performed to compare short-ANT RT between all groups while controlling for mood measures. The main effect of group when GAD-7 and PHQ-9 were included as covariates remained significant *F*(3, 122) = 7.38, *p* < 0.001, suggesting that differences in overall short-ANT RT could not be explained by differences in mood. Lastly, no significant difference in *d*’ score on the short-ANT was found in an ANOVA comparison between all groups, *F*(3, 124) = 1.77, *p* < 0.156, FDR-*p* = 0.338, η^2^ = 0.04.

Similar to the short-ANT task, ANOVA comparison of overall RT between all groups on the dot-probe task revealed a significant difference *F(*3, 124) = 14.89, *p* < 0.001, FDR-*p* = 0.004, η^2^ = 0.27. Post-hoc comparison using Tukey HSD test showed that the Pain-Meditator group (RT *M* = 409.7) performed significantly faster on all conditions in the Dot-probe task as measured by the overall RT compared to the Meditator group (RT *M* = 477.38, 95% C.I. = [9.51, 125.85], *p* = 0.018) and PainNon-Meditator group (RT *M* = 536.35, 95% C.I. = [-186.3, -67.03], *p* = <0.001). Both Meditators (95% C.I. = [13.28, 125.44], *p* = 0.01) and PainNon-Meditators (95% C.I. = [-185.94, -67.03], *p* < 0.001) also reacted slower than the Non-meditator group (RT *M* = 408.01). The remaining comparisons between groups for the overall RT on the dot-probe task did not reveal significant differences (all *p* > 0.05). The main effect between all groups remained significant in a follow-up ANCOVA performed between all groups while controlling for mood measures (GAD-7 and PHQ-9), *F*(3, 122) = 14.18, *p* < 0.001, η^2^ = 0.26).

A significant difference in *d*’ score on the dot-probe was found in an ANOVA comparison between all groups, *F*(3, 124) = 8.87, *p* < 0.001, FDR-*p* = 0.004, η^2^ = 0.18. Post-hoc comparisons using Tukey HSD test found that Pain-Meditators (*d’* = 2.88) were less accurate on the Dot-Probe than PainNon-Meditators (*d’* = 3.44, 95% C.I. = [-0.97, -0.15], *p* = 0.003), however were more accurate than Meditators (*d’* = 2.65, 95% C.I. = [-0.62, 0.18], *p* = 0.018). Meditators also performed less accurately than Non-meditators (*d’* = 3.04, 95% C.I. = [-0.78, -.004], *p* = 0.046). The remaining comparisons between groups and *d’* on the Dot-Probe did not reveal significant differences (all *p* > 0.05). See figure 1 for mean RT, *d’*, and 95% confidence interval of all groups for both the short-ANT and dot-probe tasks.

**Figure 1.**
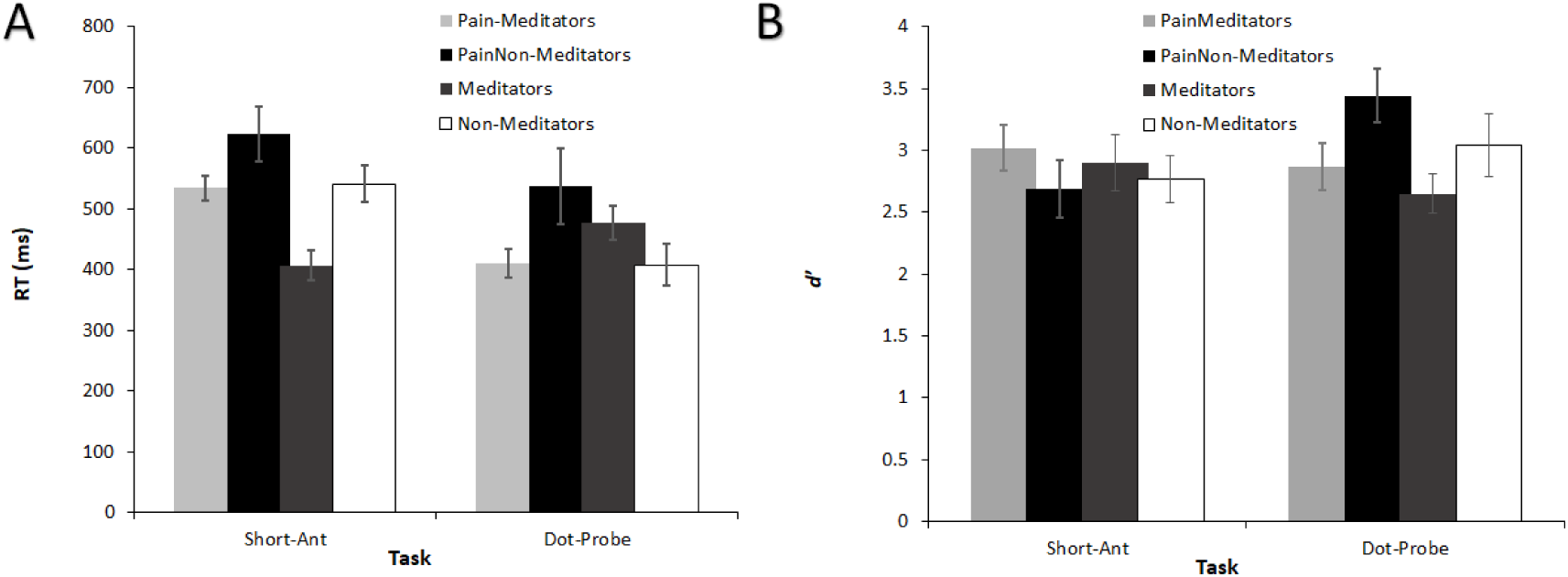
Behavioural Results for Attention Tasks: A) Average RT (in ms) for All Groups Averaged Across All Conditions for Each Attention Task, B) d’ for the All Groups Averaged Across All Conditions for Each Attention Task. Error Bars Reflect the 95% Confidence Interval of Overall Performance for Each Attention Task

### Attention network performance (Short-ANT)

Separate ANOVAs were conducted between all groups for each of the three attention network indexes (alerting, orienting, and executive control). No significant main effect between groups was found for the orienting (*F*[3, 124] = 1.30, *p* = 0.27, FDR-*p* = 0.44, η^2^ = 0.03), and executive control (*F*[3, 124] = 0.67, *p* = 0.57, FDR-*p* = 0.74, η^2^ = 0.02) networks. A significant difference in the alerting network score was found in an ANOVA comparison between all groups (*F*[3, 124] = 4.46, *p* = 0.005, FDR-*p* = 0.016, η^2^ = 0.1) (see figure 2). Post-hoc comparison using Tukey’s HSD Test found that the alerting network scores were significantly higher for PainNon-Meditators (*M* = 71.35) than Pain-Meditators (*M* = 37.01 *p* = 0.022, 95% C.I. = [-65.12, -3.55]), Meditators (*M* = 38.7, *p* = 0.031, 95% C.I. = [-63.2, -2.1]), and Non-Meditators (*M* = 33.54, *p* = 0.007, 95% C.I. = [-65.12, -3.55]), while no differences were found in comparisons between all other groups (all *p* > 0.05).

**Figure 2.**
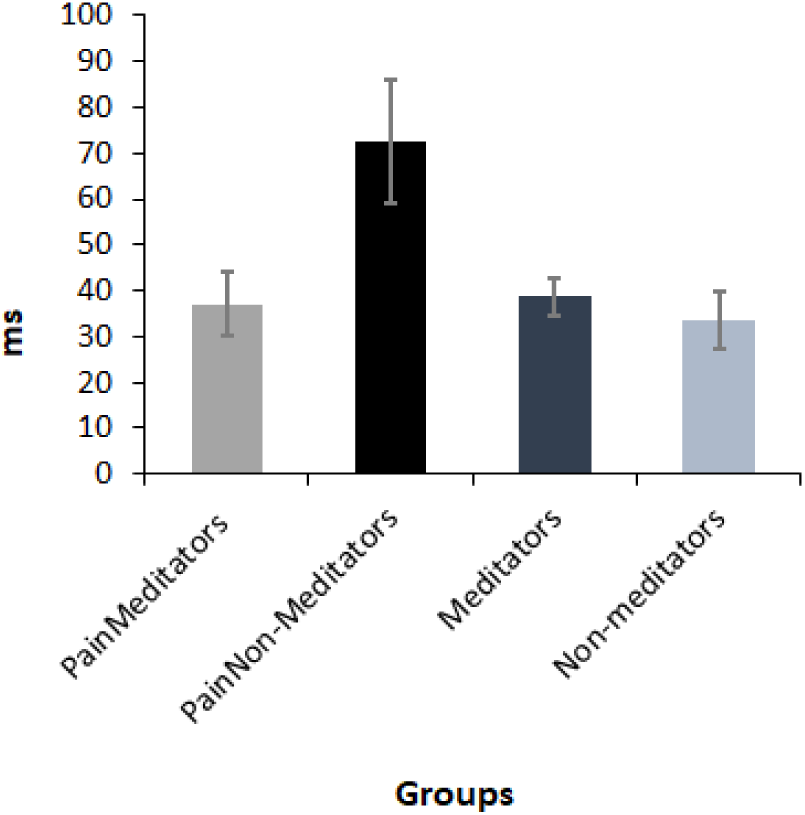
Alert Network Index Score (in ms) between Pain Groups. Error Bars Reflect the 95% Confidence Interval.

### Relationship between attention network scores and MASS, PRS, and PVAQ

To determine whether attention network scores were related to higher levels of pain resilience (PRS), mindfulness (MASS), and lower levels of pain-vigilance (PVAQ) for Pain-Meditators, a multiple regression was performed to predict attention network scores from MASS, PRS, and PVAQ. No significant results were found for all three attention network scores; alert network scores, *F*(3, 27) = 1.49, *p* = 0.239, FDR-*p* = 0.439, R^2^ = 0.14, orienting network scores, *F*(3, 27) = 0.15, *p* = 0.982, FDR-*p =* 0.982, R^2^ = 0.02, and executive control network scores *F*(3, 27) = 0.41, *p* = 0.655, FDR- *p =* 0.774, R^2^ = 0.04.

### Pain-related attention bias (Dot-probe)

To analyse the pain-related attention bias in the dot-probe task, a 2 (meditation status) × 2 (word type) × 2 (congruence) ANOVA was performed (restricted to only the pain groups). Mean RTs for each of the conditions are presented in table 2. A main effect for group was found *F*(1, 58) = 27.34, *p* < .001, FDR-*p* = .004, η^2^ = .28, however no significant interaction was found for group x word type *F*(1, 58) = .05, *p* = .824, FDR-*p* = .87, η^2^ = 3.38x10^-5^ or group x congruence *F*(1, 58) = 1.66, *p* = .203, FDR-*p* = .3, η^2^ = .001. A *t*-test was performed to compare pain-bias index scores between pain PainMeditators (*M* = 4.43, *SE* = 4.37) and PainNon-Meditators (*M* = 1.77, *SE* = 12.54), there was no effect of meditation status on pain-bias index scores, *t*(58) = 0.21, *p* = 0.84, FDR-*p* = 0.91.

**Table 2.** Average RT And Standard Error to Congruent and Incongruent Word Trials for both Pain Groups.

| Group | Location |  | *M* | *SE* |
| --- | --- | --- | --- | --- |
|  | Word Type | Probe |  |  |
| PainMeditators<br>N= 31 | **Pain Upper** | **Upper** | 397.3 | 9.09 |
|  | Pain Lower | Upper | 412.99 | 10.41 |
|  | Neutral Upper | Lower | 413.81 | 12.14 |
|  | **Neutral Lower** | **Lower** | 409.53 | 10.51 |
| PainNon-Meditators<br>N= 29 | **Pain Upper** | **Upper** | 537.84 | 29.03 |
|  | Pain Lower | Upper | 528.47 | 21.95 |
|  | Neutral Upper | Lower | 533.97 | 21.94 |
|  | **Neutral Lower** | **Lower** | 539.83 | 26.34 |
| *Note: N= number of participants; Upper= upper section of the screen, Lower= Lower half of the screen. Congruent trials are presented in bold.* |  |  |  |  |

## Discussion

The present study examined differences in attention functioning between individuals with chronic pain who have practised mindfulness and those who have not had any mindfulness experience. The results indicated that Pain-Meditators showed faster reaction times across all conditions on both the short-ANT and dot-probe attention tasks, which measured attention processing, conflict resolution, orienting, and responses to both pain-related and neutral stimuli. A higher alerting network score on the short-ANT task was found for PainNon-meditators compared to all other groups, indicating a greater difference in reaction time between the no cue and cue conditions, suggesting that perhaps PainNon-meditators benefited more from the cue to alert of upcoming trials than all other groups. Pain-Meditators did not differ in alerting network scores compared to Meditators and Non-meditators, suggesting they were perhaps less affected by pain-related attention interference than the Pain-Non-Meditators. In terms of attentional bias towards pain-related word descriptors, no differences were found between the pain groups. The results provide evidence that mindfulness practice was associated with altered attention performance in individuals suffering from chronic pain who meditate compared to individuals with chronic pain who do not meditate.

### Pain-Meditators showed faster reaction time to both attention tasks than PainNon-Meditators

As expected, Pain-Meditators demonstrated faster reaction times on both attention tasks compared to PainNon-Meditators. Although no studies specific to pain have investigated whether mindfulness training can improve attention in participants with chronic pain, the present study’s results are consistent with evidence from previous research that showed mindfulness-related improvements in attentional performance, as demonstrated by faster RTs on the ANT. For example, in a study by Becerra et al. (2017), participants in an eight-week mindfulness program showed improved ANT RT compared to waitlist controls, with shorter RTs after mindfulness intervention compared to pre-testing results. These effects have also been observed in brief mindfulness studies. Norris et al. (2018) asked participants to practice 10 minutes of mindfulness meditation using a guided mindfulness recording before completing the ANT. The authors reported that participants in the mindfulness condition performed faster on the ANT (shorter RTs) than participants in the control condition. The present study expands on these results, providing evidence that indicate mindfulness-related improvements in attentional performance are also applicable to participants with chronic pain.

The observed effect of mindfulness on attention in individuals with chronic pain may be explained by several underlying mechanisms. As previously mentioned, pain is thought to interfere with attention functioning by competing with other demands for attentional resources and therefore diminishes the individual’s ability to exert top-down control over their attention (Moore et al., 2012, 2019). However, the ability to control, orient, and maintain attention on a selected target can be trained through practices that involve attention-specific exercises (Posner et al., 2015; Tang & Posner, 2009). Mindfulness-meditation, with its focus on enhancing attentional control, typically involves attention training through the exercise of focused attention on an internal (breath or body sensation) or external (e.g., sound) stimulus while simultaneously reducing attentional focus on distractors such as thoughts, emotions, or discomforting bodily sensation (Baer et al., 2006). This process trains the practitioner’s ability to regulate their attention and is suggested to lessen the involuntary capture of attention by discomforting experiences such as pain (Baer et al., 2006; Shapiro et al., 2006). This effect has been shown in a number of studies with results consistently demonstrating an increase in both pain and discomfort tolerance after mindfulness practice (Liu et al., 2013; Mohammed et al., 2018; Shires et al., 2020). Neuroimaging research have suggested that these attention effects and changes to discomfort tolerance are the result of neuroplastic changes in mindfulness practitioners (Wang et al., 2021).

### Pain altered alerting network function, but mindfulness may protect against this alteration

PainNon-Meditators in the present study showed significantly higher alerting network scores than all other groups while no differences were found between Pain-Meditators and general population groups (Meditators and Non-meditators). The alerting network on the ANT is measured by calculating the difference in RT between the cue and no-cue conditions, and a higher score reflects a greater difference between the two conditions (Fan et al., 2002, 2005). Considering the overall slower RT on the short-ANT for PainNon-Meditators compared to other groups, higher alerting network scores could suggest that PainNon-Meditators received a greater benefit from pre-trial cues to alert of upcoming trials. Similarly, lower alerting network scores for Pain-Meditators compared to PainNon-Meditators suggests that Pain-Meditators were perhaps less reliant on pre-trial cues to signal and alert them of upcoming trials than PainNon-Meditators. Valid pre-trial cues (cues in the same location as the upcoming arrows on the short-ANT) have been shown to elicit faster responses than invalid cues (cues in the opposite location) in participants undergoing experimental pain suggesting that pretrial cues can have a positive effect on attentional processing in those experiencing pain (Gong et al., 2019).

Furthermore, the lack of difference in alerting network scores between Pain-Meditators and general population groups (Meditators and Non-meditators) points to the positive effect of mindfulness-meditation on maintaining alertness, and may be associated with upholding attention performance in individuals with chronic pain at the same level as individuals who did not suffer from chronic pain, making them equally efficient at maintaining alertness and readiness to direct attention to goal-related targets as the general population groups. The result reveals a unique effect of mindfulness-meditation on attention functioning in individuals with chronic pain – chronic pain may impair attention, but this impairment may be resolved by task relevant cues. Mindfulness may address the attention impairment, such that the cues are no longer necessary because attention is able to be allocated irrespective of cue condition. As described earlier, pain as a health-preserving mechanism competes with other goal-related stimuli for attentional resources and has been shown to inhibit the ability to divert attention away from pain, hindering alertness and attention to goal-relevant stimuli (Mazza et al., 2018; Oosterman et al., 2012). Results from the present study indicate that mindfulness training may mitigate the disruption pain has sustained attention in individuals with chronic pain, enabling more efficient processing of goal-relevant targets.

### Lack of difference in pain-bias index scores between pain-groups

Although Pain-Meditators performed faster than PainNon-meditators on the dot-probe task overall (shorter global RT), no differences were found in pain-bias index scores. This finding is not in line with our expectation that participants with chronic pain who did not meditate would show greater attentional bias to pain-related word descriptors due to increased attentional sensitivity to pain-related stimuli than participants with chronic pain who meditated (Haggman et al., 2010). Thus, while results from the present study reveal that Pain-Meditators were faster at performing the dot-probe task overall, perhaps in line with the attention network task results demonstrating a general improvement in attention, it seems that this increase in processing speed is not specific to greater efficiency at disengaging from pain-related words. There are several explanations that might account for our findings. The current literature on pain-related bias in individuals with chronic pain as measured by the dot-probe task is largely inconsistent. Although some reports have found greater attentional bias to pain-related stimuli in individuals with chronic pain compared to the general population (lower pain-index scores as measured by the dot-probe task; (Schoth et al., 2012; Todd et al., 2018), results from other studies have been less conclusive. For example, Asmundson et al. (2005) did not find group differences in pain-bias index scores between participants with and without chronic migraine headaches. Similarly, Roelofs et al. (2005) found no difference in pain-bias index scores between participants with and without chronic low-back pain using both word and picture stimuli (pictures of people performing activities that might cause back injury). Likewise, using a pictorial version of the dot-probe task, Khatibi et al. (2009) reported no difference in pain-bias scores between participants with and without a clinical diagnosis of chronic pain. As such, the task may not be sensitive to actual differences in pain bias.

The inconsistent results in the pain-bias literature using the dot-probe task are thought to be due to several factors which may explain the results of the current study (Schoth et al., 2012). For instance, the timing of stimulus presentation has been shown to impact pain-bias index results. In two separate studies by Liossi et al. (2009, 2011), the presentation of word stimuli at the 500ms timepoint after the dot-probe did not reveal group differences (pain vs no pain) in the pain-bias index, however, presentation of the stimuli at the 1250ms condition showed that participants with chronic pain demonstrated significantly greater pain-related bias than the non-pain group. The authors suggested that 500ms may not be long enough to allow for attention to orientate between threat and neutral triggers and may instead reflect the focus of attention (probe location) on the previous trial. These results could suggest that dot probe stimuli presented at the 500ms timepoint after the pain or neutral words (the procedure of the current study) may not be effective at eliciting differences in attention bias to pain-related stimuli. Furthermore, word category (affective vs sensory) can also influence pain-bias scores. Individuals with chronic pain have been shown to be more biased toward sensory words describing pain (e.g., stabbing) than affective words (e.g., sadness), and that affective word types tend to produce less group differences in pain-bias index scores between groups with and without chronic pain (Schoth et al., 2012; Todd et al., 2018). The present study did not differentiate between affective and sensory words and included both word types in the same threat category which then included both word types in the pain-bias index calculations. This lack of differentiation could therefore influence the results by conflating both affective and sensory word types.

Lastly, although no differences were found in pain-bias index scores, the PainNon-meditator group performed more accurately on the dot-probe task overall. This may be reflective of a tonic (non-stimulus specific) increase in attention to pain related cues in general (rather than an attention effect of the pain stimuli specifically as would be reflected by the pan-bias index). Also, as their RT was slower than the Pain-Meditator group, it suggests that they spent more time processing the stimuli and indicates a speed/accuracy trade off in the PainNon-meditators that was not present in the Pain-Meditator group. Perhaps the Pain-Meditator group did not have their attention automatically attracted to the pain-related cues, and so were able to respond to the task rapidly, but also did not show the performance benefits of automatic attention capture (increase in accuracy).

### Limitations and future direction

There are several limitations to the study. First, with regards to the dot-probe task, the lack of differentiation between word types (affective vs sensory) and the timing of word presentation could influence the results of pain-bias calculations. Thus, future studies investigating mindfulness-related effects on attention functioning and chronic pain could adopt a dot-probe task with randomised timing of word presentation (at 500ms and 1250ms) and include two separate conditions with affective and sensory words plainly differentiated. Furthermore, the current study included participants with meditation experience from a variety of different mindfulness-based practices. Previous research has reported unique attention network effects from different forms of mindfulness practice. For instance, Ainsworth et al. (2013) found that open monitoring and focused attention meditation techniques had an effect on the executive control network (lower network scores post-training) but not the alerting or orienting network. Baijal et al. (2011) found that concentrative meditation training improved alerting and executive control networks but not the orienting network. These results are in line with neuroimaging research that has identified unique brain activation patterns across various mindfulness practices and techniques (Fox et al., 2016).

However, previous research has noted that the difference in results between styles of mindfulness-meditation practice would likely be much smaller than differences between meditators and non-meditators, and considering the overlap between many of the different styles of mindfulness-practice, the added nuance of differences between meditation styles in attention performance may be less noteworthy than differences between meditators and non-meditators (Bailey et al., 2023). Nonetheless we acknowledge that for clinical groups, the effects of a specified set of mindfulness techniques on attention performance could facilitate more targeted treatment planning concerning pain management.

Lastly, the relatively small sample size of the present study may be underpowered, and thus may have had a lower probability of detecting the true effects of mindfulness-meditation on attention in individuals with chronic pain (Button et al., 2013; Vasileiou et al., 2018). This could explain the lack of finding in our regression analyses as smaller samples tend to have less statistical power, and a smaller sample increases the probability of both false positives and false negatives, particularly with higher numbers of predictors (Abraham & Russell, 2008; Green, 1991; Jenkins & Quintana-Ascencio, 2020). A small sample size also raises the likelihood for sampling and response biases (Burchett & Ben-Porath, 2019). Furthermore, both pain groups in the present study presented with varying conditions such as low back pain, chronic headaches, and prolonged pain sustained from various injuries. Our sample is thus a small sample of participants with varying chronic pain conditions which may affect the generalisability of the results. Individuals with different types of chronic pain conditions have been shown to have varying degrees of attention and neurocognitive function deficits which our sample was unable to account for (Crombez et al., 2013; Higgins et al., 2018).

Despite these limitations, the present study is the first to provide behavioural evidence of mindfulness-related differences in attention processing in individuals with chronic pain. The lack of difference in attention network scores between Pain-Meditators and general population groups (Meditators and Non-meditators) is particularly important for understanding the mechanism of mindfulness-related improvement in attentional functioning for chronic pain. Overall, results from this study further elucidate the effects of mindfulness on attention in individuals with chronic pain and support its potential utility as a pain-management intervention.

## Data Availability

Data in the present study are available upon reasonable request to the authors

